# SARS-CoV-2 causes a different cytokine response compared to other cytokine storm-causing respiratory viruses in severely ill patients

**DOI:** 10.1101/2020.11.14.20231878

**Authors:** Marton Olbei, Isabelle Hautefort, Dezso Modos, Agatha Treveil, Martina Poletti, Lejla Gul, Claire D Shannon-Lowe, Tamas Korcsmaros

## Abstract

Hyper-induction of pro-inflammatory cytokines, also known as a cytokine storm or cytokine release syndrome (CRS) is one of the key aspects of the currently ongoing SARS-CoV-2 pandemic. This process occurs when a large number of innate and adaptive immune cells are activated, and start producing pro-inflammatory cytokines, establishing an exacerbated feedback loop of inflammation. It is one of the factors contributing to the mortality observed with COVID-19 for a subgroup of patients. CRS is not unique to SARS-CoV-2 infection; it was prevalent in most of the major human coronavirus and influenza A subtype outbreaks of the past two decades (H5N1, SARS-CoV, MERS-CoV, H7N9). Here, we collected changing cytokine levels upon infection with the aforementioned viral pathogens through a comprehensive literature search. We analysed published patient data to highlight the conserved and unique cytokine responses caused by these viruses. A map of such responses could help specialists identify interventions that successfully alleviated CRS in different diseases and evaluate whether they could be used in COVID-19 cases.

## 1 Introduction

The current coronavirus (COVID-19) pandemic has very much focused attention upon this and other viral infectious diseases that the host antiviral immune response is unable to resolve (1–5). Indeed, major efforts are now concentrating on how severe acute respiratory syndrome β-coronavirus 2 (SARS-CoV-2) alters normal antiviral immune responses (6–15). SARS-CoV-2 causes a wide range of clinical symptoms from asymptomatic, through mild (fever, persistent cough, loss of taste and smell), to severe inflammation-driven pneumonia resulting in multiple organ failure and ultimately death (9–20). SARS-CoV2 induces an anti-inflammatory response attacking both the upper and lower respiratory tract as well as the gut. The upper respiratory tract infection could cause its high infectivity, while the lower respiratory tract and extrapulmonary symptoms are responsible for its severity. Although SARS-CoV-2 appears to modify host inflammatory defences, similar modifications are also observed in other severe respiratory infections caused by viruses such as influenza A, β-coronaviruses SARS-CoV and MERS-CoV. These agents all constitute a global health threat with colossal economic consequences (21,22).

Although these different viruses cause similar clinical symptoms, the pathogenesis may be driven by different triggers, depending upon the virus. Multiple studies have described an exacerbation of the pro-inflammatory host immune response associated with severe forms of the diseases, including cytokine storms, or cytokine release syndrome (CRS) (1–5,9–16). Although CRS usually resolves following completion of the antiviral response, it persists in severe cases leading to tissue damage, multiple organ failure and death in critically-ill patients if the clinical intervention is not rapid. In such cases, concentrations of both pro- and anti-inflammatory cytokines are significantly increased in blood and other tissues, including the type-I interferons (IFN) (IFN-□, -β, -κ, -ε, -τ, -ω and -ζ) (23– 26). Type I IFN signalling cascades also attenuate inflammation to avoid tissue damage during viral infection (27). The main effectors of the type-I IFN signalling are IFN-□ and IFN-β, which activate other cytokines such as IL-12 and the type II interferon cytokine, IFN-γ (28,29). However, cytokines such as IL-10 block the type-I IFN response. Certain pathogens, including SARS-CoV and MERS-CoV encode proteins that can influence and delay the type-I IFN response leading to various pathologies (30–32). In the case of SARS-CoV, the build-up of activated macrophages in the lungs can cause tissue damage, while MERS-CoV can intensify engagement by neutrophils, leading to an increase in the production of pro-inflammatory cytokines (33,34). Furthermore, Influenza A and coronaviruses infections can trigger increased levels of the type-I interferons IFN-□ and IFN-β, reflecting the normal initiation of this signalling pathway in response to viral infections (9–20,26,34– 38). However, in severe infections with SARS-CoV-2, the type-I IFN signalling is impaired, culminating in an altered development of adaptive immunity (9–15,35,39,40)

The broadly similar clinical symptoms and the range of disease severity of different respiratory viral infections tend to blur the accuracy of initial diagnosis (41,42), and explain why systemic non-prescription medications are given to patients to initially treat broad symptoms rather than acting on virus-specific parameters (41). Capturing a picture of the immune response triggered in each patient, early enough in infection remains challenging, impairing the adequate prevention of developing the severe form of the disease, consequently to unpredictable CRS. Defining the overlap and/or specificity in the patient immune cytokine signalling across CRS-causing viruses would help clinicians to develop a more tailored treatment strategy for future cases. Recent reviews have attempted to compare diseases caused by influenza A and β-coronaviruses (37,43–47). However, most of these studies have focused on clinical or phylogenetic parameters (virus genome, patient age, transmissibility, fatality rate, creatinine and coagulation amongst others), whilst overlooking mechanistic details relating to host immune responses. To provide mechanistic insight into the role of pro- and anti-inflammatory cytokines into the development of severe disease caused by SARS-CoV, SARS-CoV-2, MERS-CoV and influenza viruses, we need to understand the differences in cytokine responses between the different viruses.

To identify the similarities and differences in the cytokine response, we collected and analysed the patterns of cytokine changes caused by these CRS-causing respiratory viruses. By comparing available patient data from the literature, we were able to show i) where similarities lie between the immune responses mounted against these pathogens, ii) what discriminates between influenza A subtypes and coronaviruses, and iii) what are the unique aspects of the currently circulating SARS-CoV-2 virus.

## 3 Methods

### 3.1 Literature search

A mass literature search of 98 cytokines (48) was performed in PubMed using PubTator, and in bioRxiv^1^ and medRxiv^2^ non-peer reviewed pre-publication repositories (49). This included the commonly studied interleukins, interferons, tumor growth factors and chemokines involved in pro-inflammatory and anti-inflammatory responses - in particular those associated with disease-associated CRS manifestations. Only studies indicating increase or no change in cytokine levels were included. The amplitude of change was not measured, only the presence or absence of it. We focused our study on five important CRS-causing viruses: the two influenza A virus subtypes H5N1 and H7N9 and the three β-coronaviruses SARS-CoV, MERS-CoV and SARS-CoV-2 (Figure 1). We used the names of each virus and the cytokines and chemokines as search terms, e.g. “SARS-CoV-2 + CXCL10” (Figure 1). The collected studies were then screened to retain the studies using only patient-derived data, measured in at least 10 patients. A second pass was done adding “patient” to the search terms, e.g. “SARS-CoV-2 + CXCL10 + patient” in cases where the original search term yielded more than 50 hits. We only considered articles valid if they contained patient-derived data directly; cell line or model organism-based results (and reviews) were excluded. From the main text of the resulting articles, we generated a table containing the presence of the queried cytokines and their direction of change in each disease. We closed the curation on 03/06/2020 (See Supplementary Table S2 for the full list of queried cytokines). The number of discarded articles was estimated using custom python and shell scripts, available in the publication repository.

**Figure 1:**
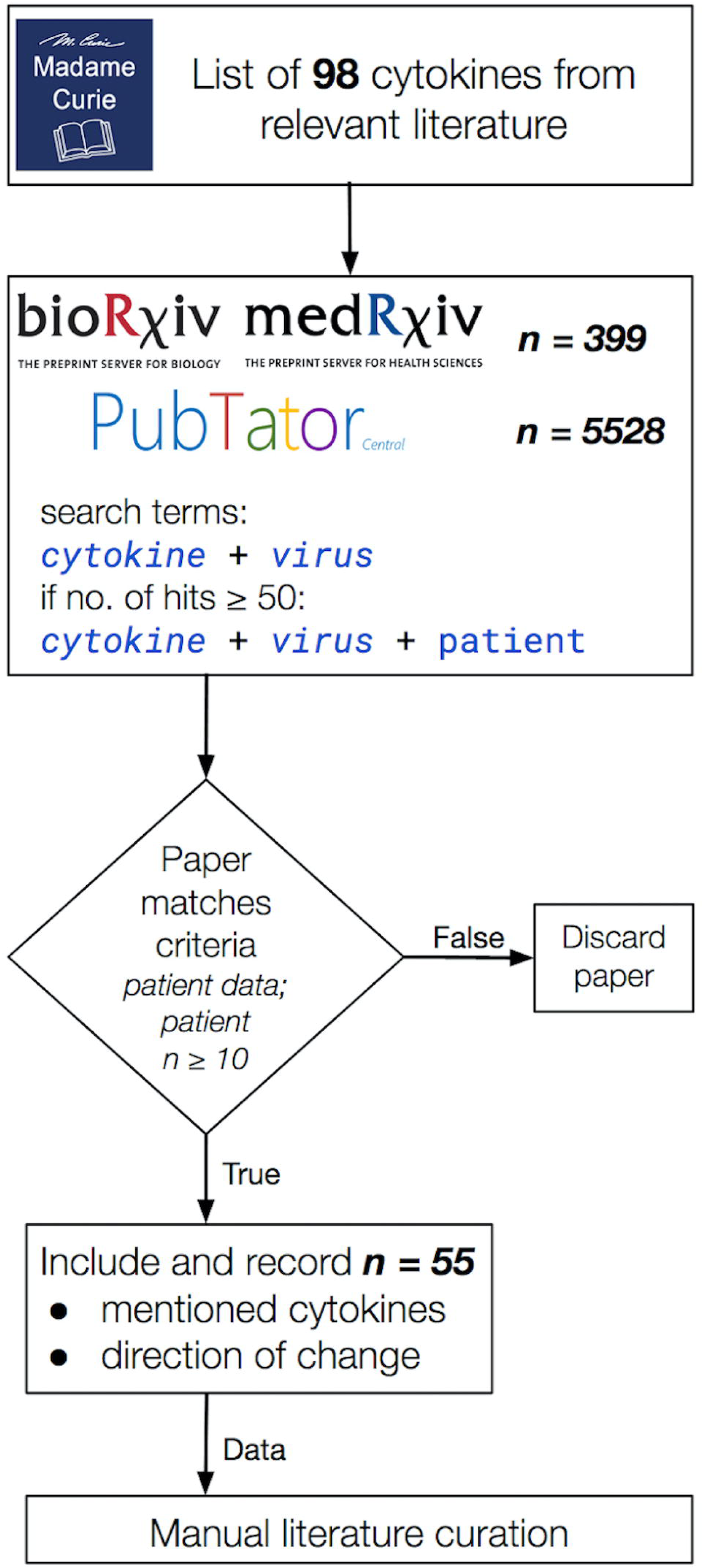
The literature curation workflow applied in this study. Publications were considered valid for inclusion into our data collection i) only if they contained patient-derived data (model organisms and cell lines were excluded) and ii) if the study data were collected from cohorts of at least 10 participants per group, iii) and if it included a change in cytokine levels. Total hits to queries in bioRxiv, medRxiv and PubTator shown separately in the second box from the top. 55 publications were selected that matched our curation criteria listed above.

### 3.2 Hierarchical clustering

We clustered our data using the *clustermap* function from the python package seaborn with Jaccard distance and complete linkage method (50). We used all cytokine categories as input. The code is available at our GitHub repository^3^.

## 4 Results

In order to capture as many studies as possible on which to apply the defined inclusion filtering for our work, we started from a list of relevant cytokines found in a textbook source (48) (Figure 1). We only used studies that reported the directional change of measured cytokines. Our curation approach allowed us to highlight shared and differing cytokine responses between influenza A and β-coronaviruses, contributing to further understanding of why SARS-CoV-2 in particular differs so much from influenza A CRS-causing viruses but also from other β-coronaviruses, also capable to inducing a cytokine storm in severe cases.

### 4.1 β-coronaviruses and influenza A viruses show marked differences in some cytokine responses

Out of the nearly 100 cytokines measured across all initially-collected studies only 38 were retained as matching our criteria (See Methods section; Supplementary Table S1). Only a small group of cytokines was commonly measured for all viruses (CXCL8, IL-6, CXCL10, IL-2, IL-10, IFN-γ, TNF-α). Across the 55 literature references used here (Figure 1), we first assessed how comparable the number of different cytokines measured in these studies was across the five CRS-causing viruses. Figure 2 shows how variable this number is between virus-specific studies (e.g. 15 for H5N1 and 26 for SARS-CoV-2). This variation probably reflects i) the increasing interest developed for CRS-causing pathologies over recent years (26 recent studies reported cytokine measurement for SARS-CoV-2 against only 10 H5N1-related studies), and ii) the increased availability and sensitivity of multiplex detection method.

**Figure 2:**
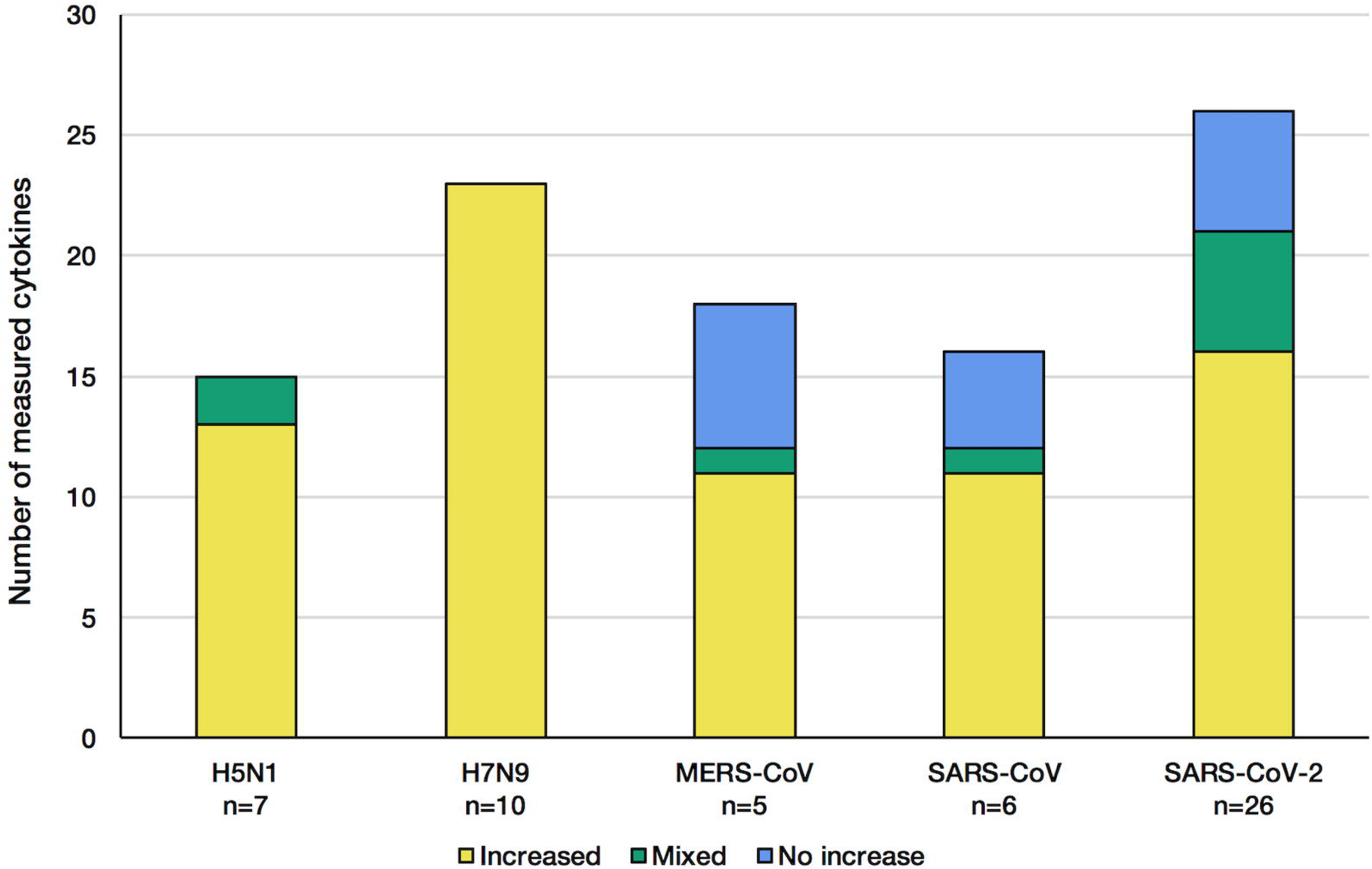
Number of cytokines measured in the studies retained for each considered CRS-causing virus. Each stacked bar indicates how many cytokines were found at increased levels (yellow) in the patients’ blood/solid tissue, not changed (blue) or both increased and not changed across different studies of the same virus (green). The *n* number shown on the bottom of the bar charts corresponds to the number of articles citing cytokine changes during infection.

The influenza A viruses trigger an increase in all cytokine levels measured (Figure 2, red). In contrast, during infection with each of the β-coronaviruses, some cytokines were detected at levels normally found in control groups (blue). This indicates that β-coronaviruses can subvert the immune response, reflecting different kinetics and pathogenesis between the influenza- and coronavirus-associated diseases. Of note, studies of H5N1 infections showed that a few cytokines were increased compared with control groups, no change was observed in other studies (17,51), illustrating the greater complexity of these diseases, probably due to the multifactorial nature of the mechanisms involved.

Table 1 shows the number of cytokines whose levels are increasing in one, or two, three, four or all five virus-related infections from the interrogated literature. Only 5 cytokines were modulated regardless of the virus-associated disease concerned, with 20 other cytokines being shared to some degree. Increased levels observed in 16 cytokines were unique to a single virus at a time. It is important to keep in mind the cytokines’ amplitude of change is not considered, which can be different between the different diseases, adding to the heterogeneity of those severe respiratory infectious diseases. This backs up the highly complex nature of the associated diseases as well as the past and current struggles to develop efficient vaccines and treatments.

**Table 1:** Number of cytokines which were elevated in at least one study. Cytokines measured in one or more of the virus-induced infections. Mixed observations occur when one or more studies show no change in a cytokine level upon infection, whereas others show an increase.

|  | Cytokines elevated at least in one study<br>(elevated & mixed) |
| --- | --- |
| Virus-specific | 16 |
| Shared between 2 viruses | 5 |
| Shared between 3 viruses | 8 |
| Shared between 4 viruses | 2 |
| Common to all 5 viruses | 5 |

To examine the presence of the measured cytokines and directionality of their change, we constructed a heatmap of the included viruses and cytokine responses.

### 4.2 The cytokine response to SARS-CoV-2 sits in between the ones given to other β-coronaviruses and influenza A viruses

The cluster analysis of the viruses forms three clusters. SARS-CoV and MERS-CoV comprise the coronavirus cluster, H5N1 and H7N9 form the influenza cluster, while SARS-CoV2 sits in an individual cluster (Figure 3), slightly closer to the two influenza A viruses than to the two β-coronaviruses.

**Figure 3.**
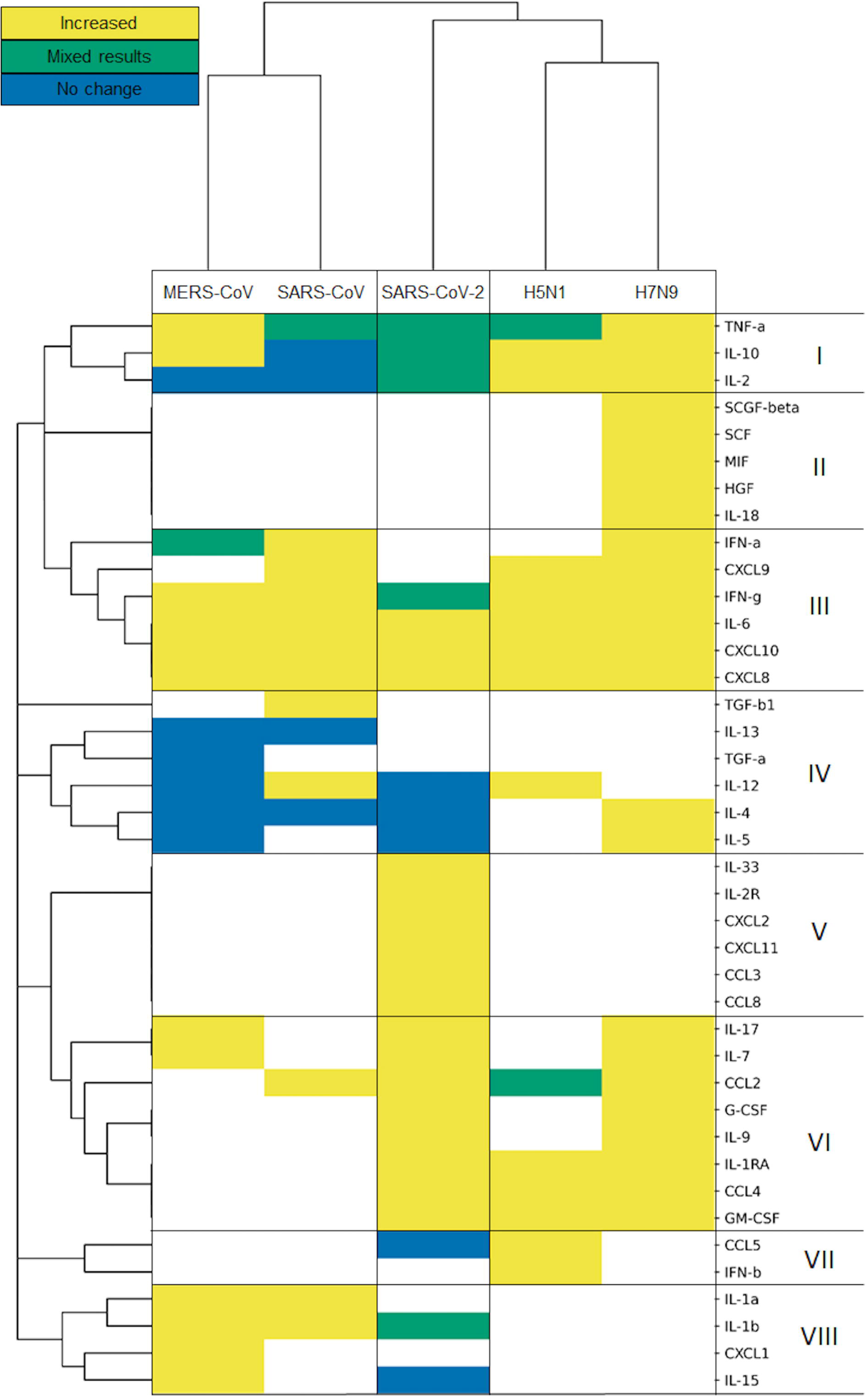
Influenza viruses, SARS-CoV and MERS-CoV, and SARS-CoV2 form separated clusters (I-VIII) based on their cytokine response. Hierarchical clustering is based on Jaccard distance complete linkage.

The cluster analysis of cytokines defines eight clusters, based on the direction of their modulation upon infection with each virus. It is important to note that the results of this cluster analysis are biased by the missing information for some cytokines. Bearing this in mind, it is nonetheless worth looking into the detailed patterns of cytokine responses of the various CRS-inducing viruses. The cytokine cluster I includes the pro-inflammatory TNF-□ and two anti-inflammatory cytokines IL-2 and IL-10. All of them had mixed results in SARS-CoV-2, while encompassing all three categories of results for the other two coronavirus infections, and predominantly increased during influenza infections. Unfortunately, cluster II seems to be restricted to cytokines measured only in H7N9-mediated infections, preventing us from comparing influenza A viruses versus with β-coronaviruses. Clusters III and VI carry the generally increased pro-inflammatory cytokines, which are elevated for almost all of the viruses, but not measured in all of the cases of cluster VI. Among those cytokines are IFN-□ and IFN-γ, typical representatives of the general antiviral response (type I and type II interferons), as well as IL-6, one of the most prominent pro-inflammatory cytokines, along various chemokines. Cytokines from Cluster IV measured during coronavirus infections do not fluctuate, while most of them are elevated during influenza infection, e.g. IL-4 and IL-5 upon H7N9 infections. IL-4 is involved in Th2 differentiation, and the Th2 cells can produce IL-5 to mitigate eosinophil infiltration (52). Such differences observed between virus-specific pathologies reflect the strong alterations observed in coronavirus infections, particularly SARS-CoV-2 (53). The cytokines in Cluster VII and VIII do not always respond to SARS-CoV-2: IL-15 and CCL5 (RANTES) are not elevated after SARS-CoV2 infection. IL-15 is involved in natural killer cell differentiation as part of antiviral response (54). Meanwhile, CCL5 mediates eosinophil infiltration which is considered to be involved in the recovery after SARS-CoV infection (55). Clusters II and V contain cytokines measured only in H7N9 and SARS-CoV2, respectively, whereas TGF-b1 was measured only in SARS-CoV studies in cluster IV.

For a subset of pro-inflammatory cytokines, the clustering analysis shows that SARS-CoV-2 induces a similar response to all other viruses (See Cluster III and VI; Figure 3). Its uniqueness lies in the fact that not all of the expected cytokines are elevated, e.g. the ones following an influenza infection such as IL-2, IL-10, TNF-□, IL-4 or IL-5.

### 4.3 Type I interferon signalling is more strongly altered upon infection with SARS-CoV-2 than in SARS-CoV- or MERS-CoV-infections

Both type-I and type-II IFNs play an instrumental role in the immune response to viral infection.

Our analysis shows early induction of type-I interferons occurs upon H5N1 and H7N9 influenza A infection as well as for the β-coronavirus SARS-CoV and MERS-CoV (26,37,56). However, type-I IFN response is only weakly elicited following a SARS-CoV-2 infection, if at all (39,57, Figure 4).

**Figure 4:**
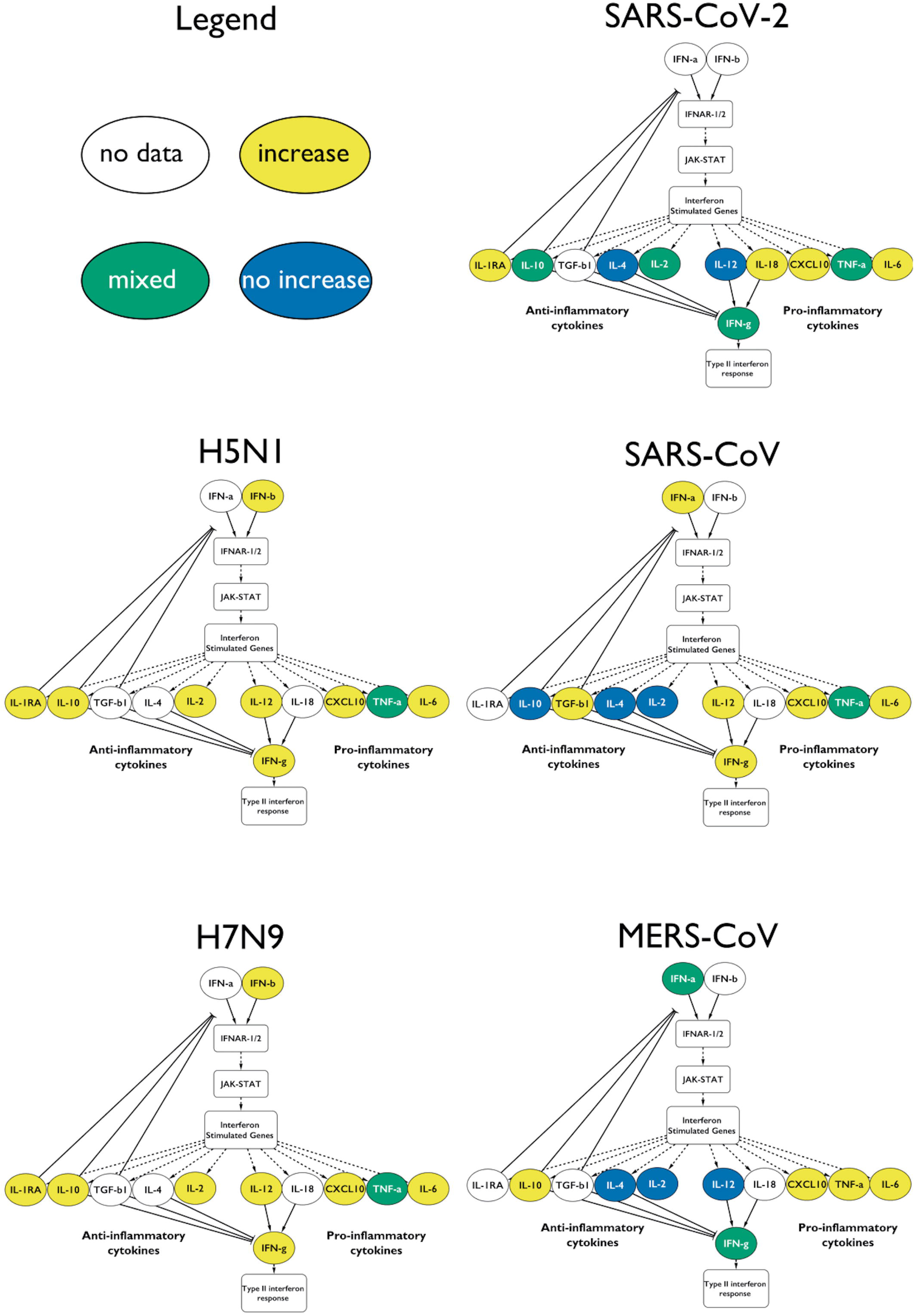
Type I interferon response upon infection with the different CRS-causing viruses. The measured cytokines in the influenza viruses are increased. In the case of the coronaviruses the response is mixed, not all of the anti-inflammatory cytokines are elevated. Only a fraction of cytokines is depicted for clarity. Yellow for increase in that virus, green for mixed results and blue for no-change.

Infection with either of the two influenza subtypes seems to increase the levels of measured type-I IFN-relevant cytokines, resulting in an antiviral immune response, with the appropriate cytokines showing elevated levels in all influenza A studies (Figure 4, Supplementary Table S1).

The β-coronavirus-mediated responses show a much more variable IFN response: with SARS-CoV, we see that the type-I IFN response is active, including the downstream-activated IL-12 that reflects the involvement of mature dendritic cells. IL-12 also indirectly activates IFN-γ further downstream. IL-10 is not elevated, which potentially prevents the downregulation of the type-I interferon response.

In MERS-CoV infections, the type-I IFN response is induced, but not in all cases (58). In some studies, the levels of IL-12 do not increase, in agreement with IFN-γ also staying at low levels. Yet we see the involvement of the (mostly) anti-inflammatory IL-10. However, caution needs to be applied when looking at IL-10 in an inflammation context, as more and more clinical evidence suggests this cytokine displays pro-inflammatory characteristics *in vivo* (59,60).

We showed here that SARS-CoV-2-mediated infections are characterized by a clear dysregulation of type-I IFN response, and consequently the downstream cytokine signature such as IL-4, IL-12, IL-2, IL-10 and the downstream type II IFN response (Figure 4).

## 5 Discussion

In this study, we analysed relevant cytokine levels measured in patients infected each with one of five major viral pathogens through a comprehensive literature curation of published patient data. We generated a map of such responses to help specialists identify routes of interventions to successfully alleviate CRS in different diseases, and evaluate whether they could be used in COVID-19 cases. Based on our literature curation, the five investigated viruses cause atypical cytokine responses in severely-ill patients, reported here in Figure 3.

The cytokine response during viral infection is a dynamic process, with multiple changes in cytokine levels during the course of the infection. During SARS and MERS infection, a slow initial innate immune response accompanied by the infection of alveolar macrophages leads to increased severity of these lower respiratory tract diseases (61–65). Furthermore, a long-lasting pro-inflammatory cytokine production results in high mortality due to the development of severe conditions such as acute respiratory distress syndrome (ARDS) or acute lung injury (9.5% fatality rate for SARS and 34.4% for MERS compared to 2.3% for COVID-19 (44)).

Severe SARS patients show particularly low levels of the anti-inflammatory cytokine IL-10 (Figures 3, 4) (66). During MERS infection patients develop an expected increased production of IL-10, yet the low levels of IFN-γ-inhibiting IL-4 and IL-2, lead to elevated IFN-γ and the induction of type-II IFN response (Figure 3) (58,67,68). In contrast, during influenza A infection, the antiviral response activates without much delay with the presence of an intact negative feedback loop. Both viruses considered in our curation induce most of the pro- and anti-inflammatory cytokines downstream of type-I interferon response (Figure 3). Although influenza A viruses have effectors that dysregulate IFN-I (e.g. NS1, PB1-F2, polymerase proteins), the IFN-I response is nonetheless sustained, and its excessive activation during severe illness can lead to increased mortality. Furthermore, during H7N9 and H5N1 severe infections, TGF-β fails to be activated, contributing to increased pathogenicity (69– 71). SARS-CoV-2 stands out from the other β-coronaviruses and influenza A viruses, with a highly perturbed response downstream of Type I IFN signalling, as reflected in the poor balance of measured pro-and anti-inflammatory cytokines (Figures 3 and 4). Of note, IFN-□ was found to be increased (similar to the other viruses) only in one small (n=4) patient study, which did not match our inclusion criteria. Type-II IFN-γ was also only increased in patients placed in intensive care units (ICUs), while it was within normal ranges in other studies (10,72,73).

Although the cytokine signalling enabling the reduction of the inflammatory environment is active (Figures 3 and 4), both influenza viruses H5N1 and H7N9 can cause CRS. In severe cases of infection, CRS could result from insufficient production of important cytokines such as TGF-β (71). Furthermore, the presence of impaired and less abundant effector CD4+ and CD8+ T cells was found to be a characteristic feature accompanying CRS in those diseases. Finally, monocytes, that normally would differentiate from a pro- to anti-inflammatory state with enhanced antigen presentation activity as the infection progresses, remain in a chronic pro-inflammatory activation state, preventing the normal resolution of the host response (16,74,75). In future studies, patient-derived data including the size and activation status of innate and adaptive immune cell populations would help increase the understanding of CRS mechanisms in influenza-mediated diseases.

In our study, we found resolution of the pro-inflammatory immune response to be a key difference between coronaviruses (MERS-CoV and SARS-CoV) and influenza viruses (H5N1 and H7N9). Both MERS-CoV and SARS-CoV induce CRS, yet they also appear to impair the normal resolution of the antiviral immune response. In contrast, H5N1 and H7N9 induce high levels of pro- and anti-inflammatory cytokine levels in severe cases leading to an inflammatory cytokines storm, yet leaving the immune system unimpeded to move towards a general a resolution of the antiviral response appears (Figures 3 and 4) (17). However, SARS-CoV-2 induction of the CRS is eventually followed by a resolution of the pro-inflammatory responses in 80% of the cases.

One limitation of this study is the lack of anatomical and dynamic dimensions of the cytokine response. Firstly, the set of cytokines measured in the peripheral blood of each patient across the entire disease course or following recovery varied across the studies analysed. Patients were sampled at different stages of the disease, which further adds to the noise observed in the data. Finally, systematic patient-based studies matching our strict curation criteria could not be collected, leaving many gaps in our comparisons (Figure 3, white cells).

While confirming many already reported disease traits, our analysis has highlighted several new features that are shared or different between the viral diseases analysed, contributing to filling the gap in the understanding of SARS-CoV2 and other CRS-causing viruses. Blockage of the cytokine response in SARS-CoV2 infection through IL-6 specific antibody has failed during Phase 3 randomised clinical trial (NCT04320615), even with promising results in earlier stages (76), suggesting that further mechanistic investigation of the cytokine storms during SARS-CoV-2 infection will be needed.

The ongoing accumulation of patient-derived large data sets will inform the research community and clinicians of the intricacy of host/virus interactions (77). Here we provided a literature curation of patient-derived data and a comparative map across CRS-causing β-coronaviruses and influenza A viruses, linking shared or specific changing cytokines and interferon signalling alterations to those pathogens.

## Data Availability

All associated data can be found in the supplementary tables, and on the github page linked in the manuscript, in the methods section.

## Acknowledgements

We thank the current and past members of the Korcsmaros group and the COVID-19 Disease Map Community for their ideas and support.

## Footnotes

1 https://www.biorxiv.org/

2 https://www.medrxiv.org/

3 https://github.com/NetBiol/Covid19/tree/master/CRS

